# PSYCHOLOGICAL DISTRESS OF CHILDREN WITH SICKLE CELL DISEASE IN MULAGO NATIONAL REFERRAL HOSPITAL, KAMPALA, UGANDA

**DOI:** 10.1101/2025.04.30.25326713

**Authors:** Geoffrey Odong, Gerald Humble Apule, Ronald Ameny, David Nsubuga Mayanja, Emmanuel Kiberu, Natasha Kabasere, Solin Lutada Lakareber, Derrick Bary Abila, Felix Bongomin, Sarah kiguli

## Abstract

**Background:** Children living with sickle cell disease (SCD) experience multiple challenges that impairs their physical, psychological, social life and development. There is limited literature on the prevalence rate of psychological symptoms among these children in Uganda. We aimed to assess the health-related psychological distress and associated factors among children with SCD in an urban tertiary health facility in Uganda

**Methods:** A descriptive, cross-sectional study was conducted in Mulago National Referral Hospital (MNRH) Kampala, Uganda, including children with SCD aged 7 to 17 years attending inpatient and outpatient departments of sickle cell unit. Psychological distress was assessed using distress questionnaire 5 (DQ-5). We also used the Child behavioral questionnaire in a Paper based form for data collection.

**Results:** A total of 288 children were included in this study. Their mean age was 12.0±3.6 years. Majority of the children were female (52.4%, n=151), in primary schools (66.3%, n=191) and lived with their mothers only (51.0%, n=147). The prevalence of psychological distress was 34.0% (n=98). Children aged 11 to 17 years were more likely to have psychological distress when compared to those aged 7 to 10 years [Adjusted Odd Ration (aOR): 1.8, 95% Confidence Interval (CI): 1.1 – 3.1, p= 0.029]

**Conclusions:** Almost every 1 in 3 children with SCD in Uganda has psychological distress. There is a great need for interventions to support the psychological needs of this population of children. Therefore, by fostering understanding and providing appropriate support, it is possible to mitigate the negative psychological impact and promote resilience in these young patients.

## INTRODUCTION

Sickle cell disease (SCD) refers to a group of inherited hemoglobin disorders characterized by a predominance of abnormal sickle hemoglobin (HbS) in erythrocytes and is the most common genetic hematological disorder. Sickle cell anemia (SCA), which results from homozygous inheritance of sickle hemoglobin from both parents is the most common and severe form of sickle cell disease(1). It has been acknowledged as a global public health threat with significant impact by the World Health Organization (WHO) (2). Approximately 400,000 babies with SCD are born each year worldwide including over 300, 000 with SCA with the greatest burden in Sub-Saharan Africa where 75% of all SCD occurs with an expected increase by 2050 (3). Such HbS individuals usually die from an infection or severe anemia, those who survive into adulthood remain susceptible to exacerbations of the disease and its medical and psychosocial complications (4).

SCD affects close to 200,000 Ugandans many of whom lack access to adequate clinical management services and as such are frequently plagued by complications like recurrent Vaso-occlusive crises and infections (5). Currently, about 33,000 babies are born annually with the disease in Uganda of which 80% die before the age of 5. With the present lack of cure, many people with SCD are believed to live in fear of early death or have death anxiety and many other psychological complications (6).The psychological issues for people with SCD and their families mainly results from the impact of pain and symptoms on their daily life, the society’s attitude to SCD and those affected.

Sickle cell anemia is a troublesome disease for both patients and their families. Frequent emergency department visits and hospitalizations to manage pain and other complications, chronic medical treatment, inadequate nutrition, and economical and psychological problems of patients’ families are the principal factors for the development of psychiatric disorders in patients with SCA (6). In the same vein, several studies have shown that the frequency of neuropsychiatric problems in children with SCD is higher than that in normal controls with depression, anxiety disorders, learning problems, and attention deficit being the most reported (7). For children, challenges vary across developmental stages and include reduced participation in recreational activities, high levels of school absenteeism, delayed puberty, limitations in social functioning, and overall low quality of life (8). The implications are that psychological symptoms are known to contribute to Vaso-occlusive crisis and other physical complaints (9). Patients with the most clinically severe pain also show the greatest prevalence of depression (10).

Although it is a challenge determining the exact prevalence rate of a psychological disorder in any given population, some countries have attempted it and have some figures that guide action, policy, and research. In Uganda, the previous studies assessing the psychosocial burden of SCD on caretakers and the prevalence of depressive disorders within SCD have been done but only with a focus on adults and specific to depressive disorders. A 2020 study in Uganda showed a uniquely high proportion (68.2%) of adult patients with SCD suffering from depression (11) which was partly attributable to the effects of a lower socioeconomic environment which has been shown to increase the risk of depression (12).

The psychological burden in children with SCD in Uganda is substantial but poorly researched with no records of national statistics. Therefore, this study aimed at assessing the health-related psychological distress and associated factors among children with sickle cell disease in Mulago National referral hospital, Kampala, Uganda.

## METHODS

### Study area

The study was conducted in Mulago National Referral Hospital, Kampala City, Uganda between May and July 2023.

### Study design

We conducted a descriptive cross-sectional study among children with Sickle cell disease. We used quantitative approach and simple systemic sampling technique.

### Study population

We evaluated children aged 7-17 and included both the inpatient and those in the outpatient department attendees whose next of kin provided written informed assent.

### Sample size

A sample size of 288 was estimated using the **Kish Leslie formula. T**his was on assumption of a Z value of **1.96** (at cut-off level **α of 0.05**); an estimated proportion of children psychologically distressed of **0.75**, estimated proportion of children not psychologically distressed of **0.25** and we entertained error of **+/-0.05** between our “estimated” and the “true” proportions of children with psychological distress.

### Sampling techniques

In this study, simple random sampling method was used to determine the patient to interview.

### Data collection techniques

Psychological distress was measured using distress questionnaire 5 (DQ-5). It is a comprehensive questionnaire composed of different sections. The first section included the socio-demographic characteristics (Factors associated with distress among children with Sickle cell disease) like age, Sex, Level of education, Birth order, Person with whom the child resides (current custody). The second part of the questionnaire was the psychosocial effect of sickle cell disease on children, this includes five questions, “My worries overwhelmed me, I felt hopeless, I found social settings upsetting, I had trouble staying focused on tasks, Anxiety or fear interfered with my ability to do the things I needed to do at school or at home”. We also used the Child behavioral questionnaire which assessed the behavior of the children. Secondary data was collected by reviewing documentary source [Records].

### Data processing and analysis

Data was processed using Kobo Collect tool and statistical analysis using STATA 16 software. Descriptive statistics were used to summarize Socio-demographic variables. All categorical variables were summarized as frequencies and proportions.

Batterham in his study set standard cutoff points to classify patients into different level of current psychological distress. We used value 14 above which patient was psychologically distressed. This value is more specific, sensitive and reduces the rate of false positive during the study (13)

We used bi-variable and multi-variable analysis (Using poison regression analysis) to determine the association between demographic characteristics and psychological distress. We reported Crudes odds ratios, adjusted odds ratios, P-values and 95% confidence intervals.

### Ethical consideration

Our study received an ethical approval from Mulago National Referral Hospital Institutional Review Board under reference number MHREC 2425 on 15^th^ February 2023 to run for a period of one (1) year. It solely utilized deidentified information. All activities were performed in line with relevant guidelines and regulations from Mulago Hospital Research and Ethics Committee (MHREC). Written assent was obtained from the child and consent from parent or guardian. Only children who assented to the study and consent from the parents or guardian were included and reverse is true therefore, informed consent was obtained from all subjects and their legal guardians.

A unique identifier (number) was used for the purpose of confidentiality. Participating in the study was strictly voluntary in nature.

## RESULTS AND DISCUSSIONS

### Socio-demographic characteristics

A total of 288 children living with SCD were included in this study. The mean age of the children was 12.0 years (standard deviation, S.D of 3.6) with a median of 12 years. Majority of the children were aged 11 to 17 years (64.2%), were female (52.4%), were in primary school (66.3%), and lived with a mother only (51.0%) (**Table 1**).

**Table 1:**
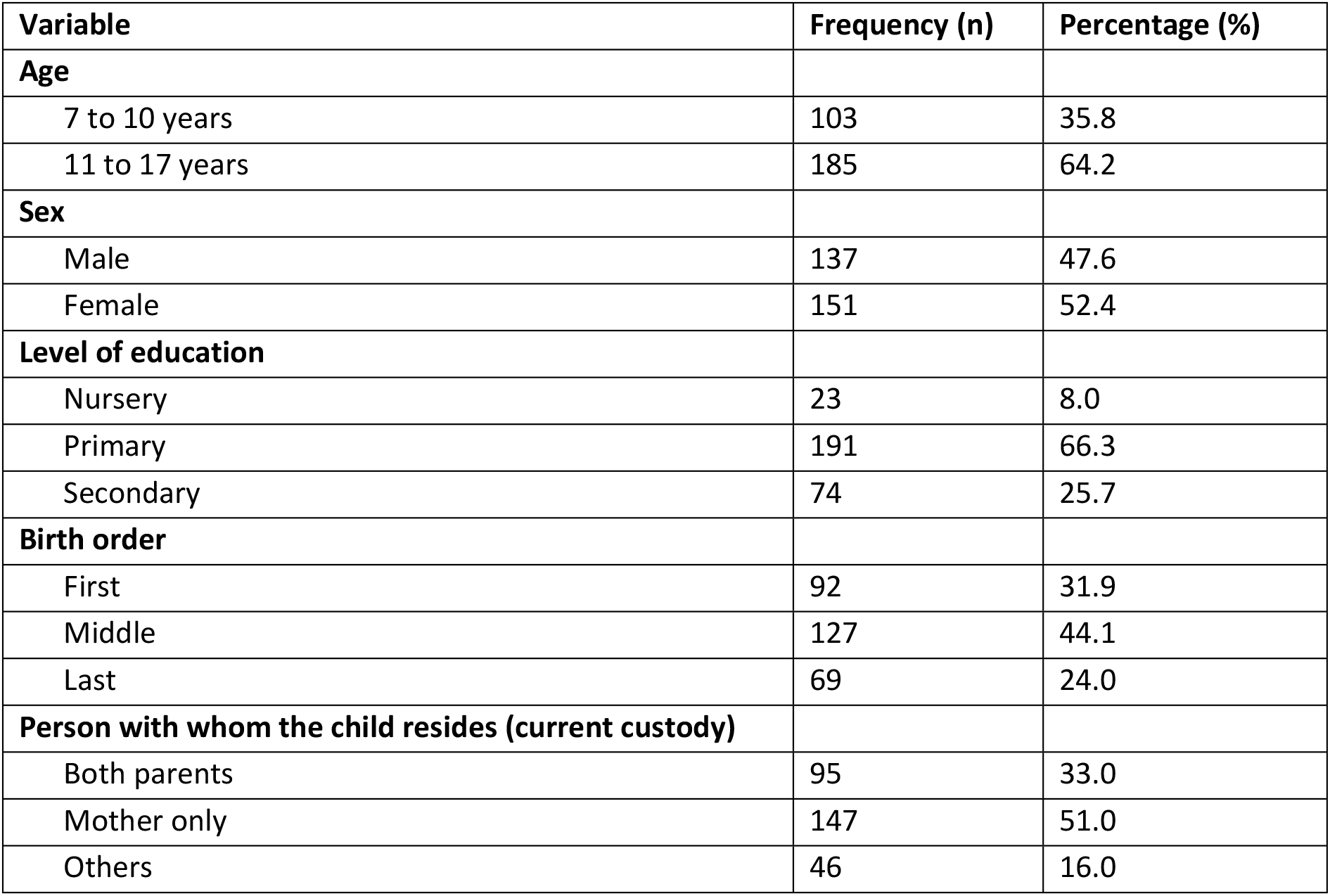
Socio-demographic characteristics of the study participants.

| **Variable** | **Frequency (n)** | **Percentage (%)** |
| --- | --- | --- |
| **Age** |  |  |
| 7 to 10 years | 103 | 35.8 |
| 11 to 17 years | 185 | 64.2 |
| **Sex** |  |  |
| Male | 137 | 47.6 |
| Female | 151 | 52.4 |
| **Level of education** |  |  |
| Nursery | 23 | 8.0 |
| Primary | 191 | 66.3 |
| Secondary | 74 | 25.7 |
| **Birth order** |  |  |
| First | 92 | 31.9 |
| Middle | 127 | 44.1 |
| Last | 69 | 24.0 |
| **Person with whom the child resides (current custody)** |  |  |
| Both parents | 95 | 33.0 |
| Mother only | 147 | 51.0 |
| Others | 46 | 16.0 |

### Psychosocial effects of Sickle cell disease on the children

The prevalence of psychological distress among the children living with SCD was 34.0% as noted in **Table 3**. Batterham in his study set standard cutoff points to classify patients into different level of current psychological distress. We used value 14 above which patient was psychologically distressed. This value is more specific, sensitive and reduces the rate of false positive during the study (13) **Table 2** summarizes the findings from the distress questionnaire 5 that examined the psychological distress among children living with SCD.

**Table 2:**
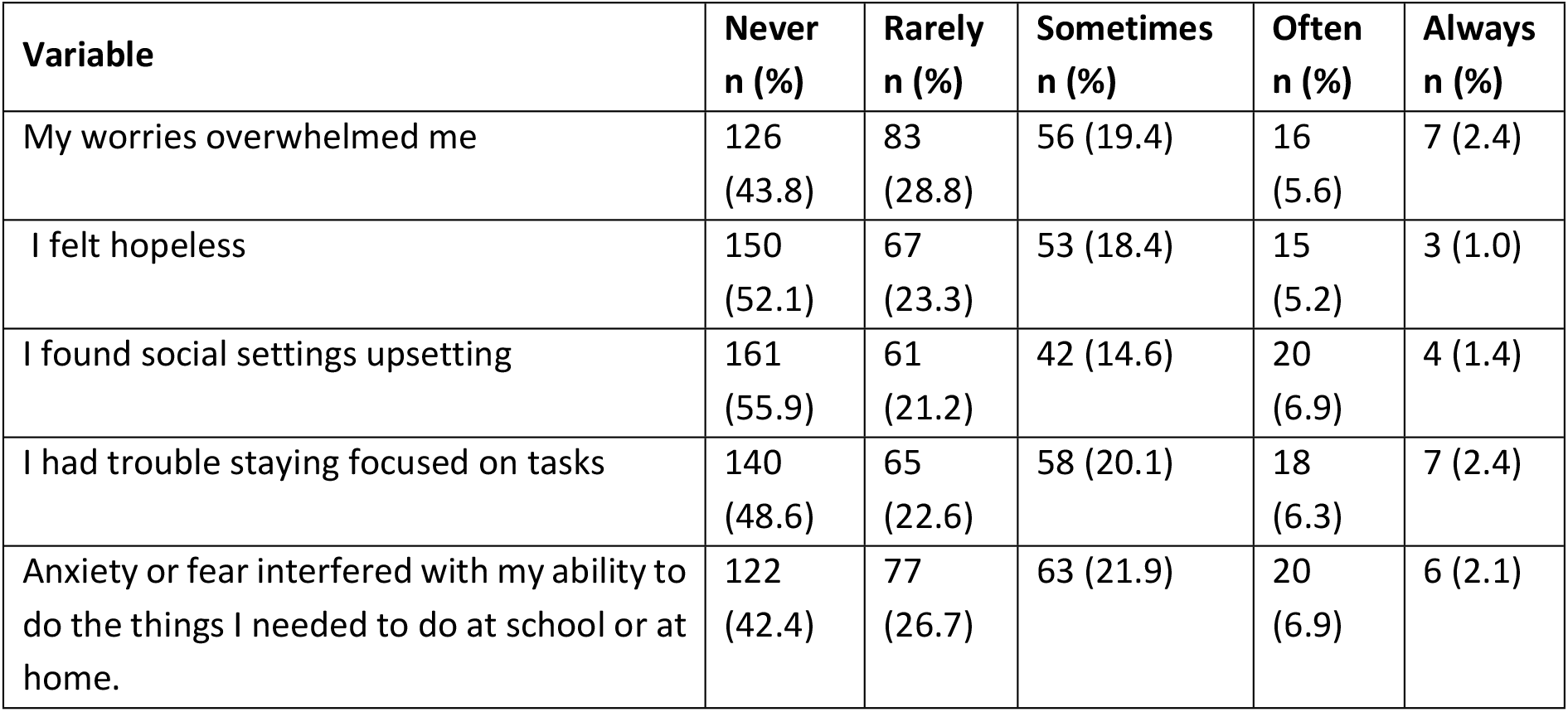
Psychosocial effects of Sickle cell disease on the children.

**Table 3:** Psychological distress among children with SCD.

| Psychological distress | Frequency (n) | Percentage (%) |
| --- | --- | --- |
| Not distressed | 190 | 66.0 |
| Distressed | 98 | 34.0 |

### Child behavior

The children missed an average of 9.5 (S.D of 10.1) days per term with a median of 7.0 days. The children had an average of 3.4 (S.D of 2.5) hospital visits and had an average of 3.0 (3.5) previous admissions in the past 12 months. Majority of the children felt that SCD interfered with their daily play activities (70.8%) and interfered with their domestic activities (56.3%). Also, majority did not feel that they were a burden to their families (62.5%), did not feel discriminated against (68.4%) and did not feel that they had bad luck (62.5%) (**Table 4**).

**Table 4:**
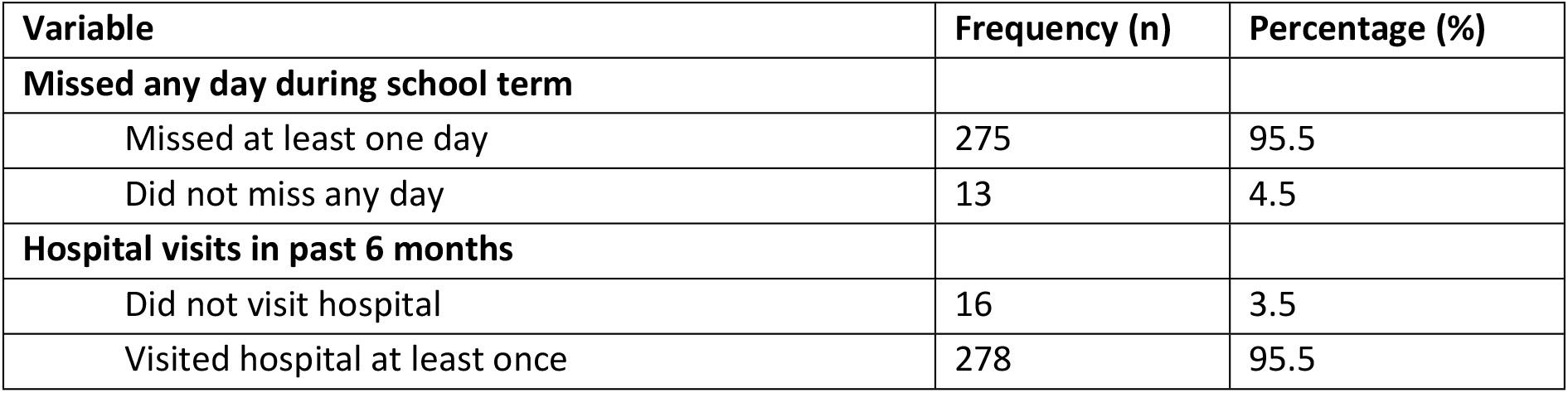
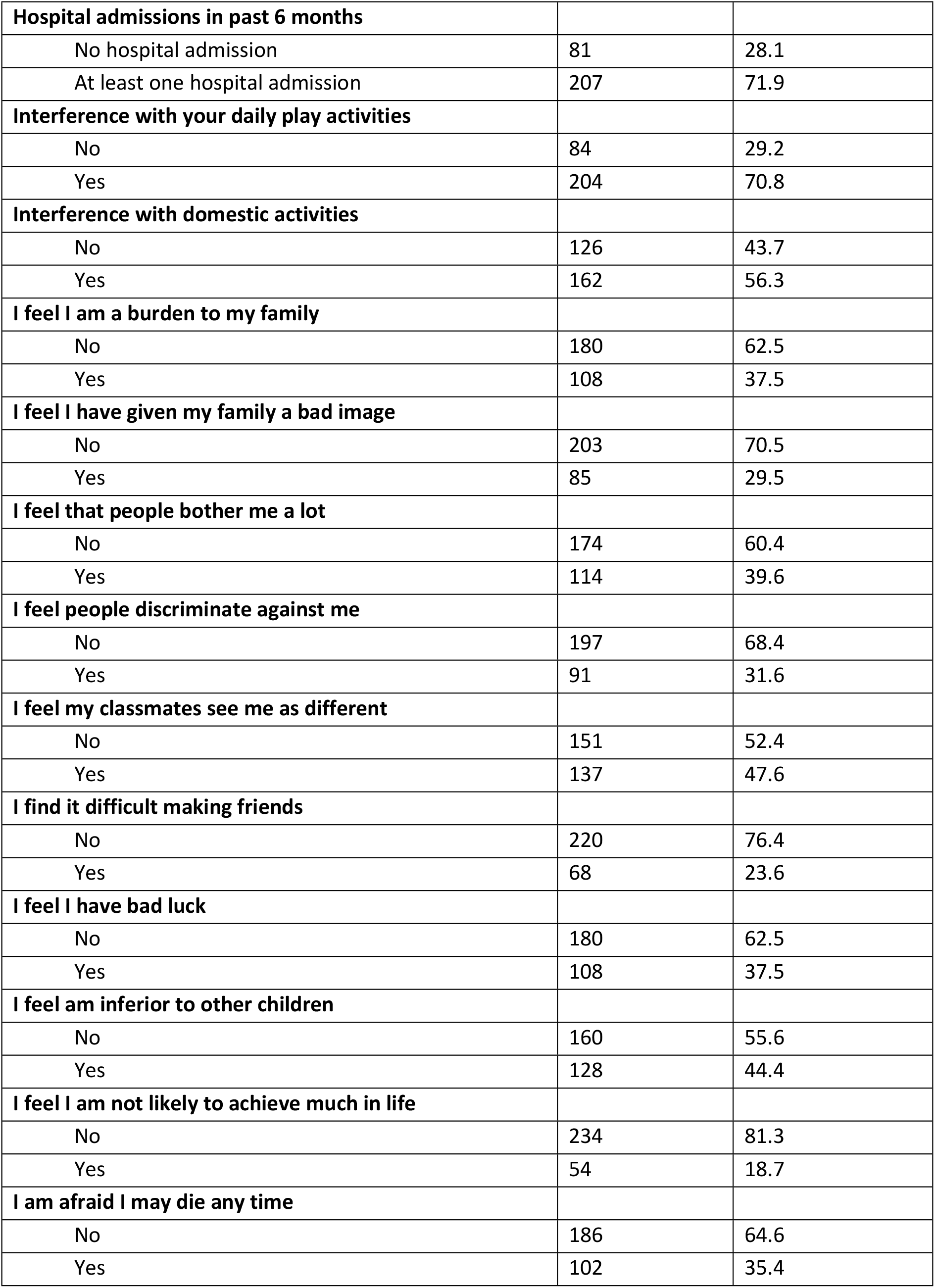
Behavior of the children.

| Variable | Frequency (n) | Percentage (%) |
| --- | --- | --- |
| **Missed any day during school term** |  |  |
| Missed at least one day | 275 | 95.5 |
| Did not miss any day | 13 | 4.5 |
| **Hospital visits in past 6 months** |  |  |
| Did not visit hospital | 16 | 3.5 |
| Visited hospital at least once | 278 | 95.5 |
| **Hospital admissions in past 6 months** |  |  |
| No hospital admission | 81 | 28.1 |
| At least one hospital admission | 207 | 71.9 |
| **Interference with your daily play activities** |  |  |
| No | 84 | 29.2 |
| Yes | 204 | 70.8 |
| **Interference with domestic activities** |  |  |
| No | 126 | 43.7 |
| Yes | 162 | 56.3 |
| **I feel I am a burden to my family** |  |  |
| No | 180 | 62.5 |
| Yes | 108 | 37.5 |
| **I feel I have given my family a bad image** |  |  |
| No | 203 | 70.5 |
| Yes | 85 | 29.5 |
| **I feel that people bother me a lot** |  |  |
| No | 174 | 60.4 |
| Yes | 114 | 39.6 |
| **I feel people discriminate against me** |  |  |
| No | 197 | 68.4 |
| Yes | 91 | 31.6 |
| **I feel my classmates see me as different** |  |  |
| No | 151 | 52.4 |
| Yes | 137 | 47.6 |
| **I find it difficult making friends** |  |  |
| No | 220 | 76.4 |
| Yes | 68 | 23.6 |
| **I feel I have bad luck** |  |  |
| No | 180 | 62.5 |
| Yes | 108 | 37.5 |
| **I feel am inferior to other children** |  |  |
| No | 160 | 55.6 |
| Yes | 128 | 44.4 |
| **I feel I am not likely to achieve much in life** |  |  |
| No | 234 | 81.3 |
| Yes | 54 | 18.7 |
| **I am afraid I may die any time** |  |  |
| No | 186 | 64.6 |
| Yes | 102 | 35.4 |

### Factors associated with distress among children with SCD

Psychological distress in children with SCD is associated with socio-demographic, clinical, therapeutic, and evolutionary factors which vary across studies. In this study, the main factor found to be associated with psychological distress in children with SCD is age (**Table 5**)

**Table 5:** Factor associated with psychological distress among children with SCD.

| Variable | Not distressed<br>n (%) | Distressed<br>n (%) | P-value |
| --- | --- | --- | --- |
| **Age** |  |  |  |
| 7 to 10 years | 77 (74.8) | 26 (25.2) | 0.019 |
| 11 to 17 years | 113 (61.1) | 72 (38.9) |  |
| **Sex** |  |  |  |
| Male | 85 (62.0) | 52 (38.0) | 0.180 |
| Female | 105 (69.5) | 46 (30.5) |  |
| **Level of education** |  |  |  |
| Nursery | 17 (73.9) | 6 (26.1) | 0.217 |
| Primary | 130 (68.1) | 61 (31.9) |  |
| Secondary | 43 (58.1) | 31 (41.9) |  |
| **Birth order** |  |  |  |
| First | 60 (65.2) | 32 (34.8) | 0.402 |
| Middle | 80 (63.0) | 47 (37.0) |  |
| Last | 50 (72.5) | 19 (27.5) |  |
| **Person with whom the child resides (current custody)** |  |  |  |
| Both parents | 61 (64.2) | 34 (35.8) | 0.111 |
| Mother only | 104 (70.8) | 43 (29.2) |  |
| Others | 25 (54.4) | 21 (46.6) |  |
| **Missed any day during school term** |  |  |  |
| No | 11 (84.6) | 2 (15.4) | 0.147 |
| Yes | 179 (65.1) | 96 (34.9) |  |
| **Hospital visits in past 6 months** |  |  |  |
| Did not visit hospital | 7 (70.0) | 3 (30.0) | 0.784 |
| Visited hospital at least once | 183 (65.8) | 95 (34.2) |  |
| **Hospital admissions in past 6 months** |  |  |  |
| No hospital admission | 60 (74.1) | 21 (25.9) | 0.069 |
| At least one hospital admission | 130 (62.8) | 77 (37.2) |  |

From the multi-variable analysis, children who were aged 11 to 17 years were more likely to have psychological distress when compared to those aged 7 to 10 years [Adjusted Odd Ration (aOR): 1.8, 95% Confidence Interval (CI): 1.1 – 3.1, p-value = 0.029] (**Table 6**).

**Table 6:** Bivariable and multi-variable analysis of factor associated with psychological distress among children with SCD (Using Poisson regression analysis).

| Variable | Bivariable analysis |  | Multi-variable analysis |  |
| --- | --- | --- | --- | --- |
|  | cOR | p-value | aOR | p-value |
| **Age** |  |  |  |  |
| 7 to 10 years | 1 |  | 1 |  |
| 11 to 17 years | 1.9 (1.1 – 3.2) | **0.021** | 1.8 (1.1 – 3.1) | **0.029** |
| **Sex** |  |  |  |  |
| Male | 1 |  | 1 |  |
| Female | 0.7 (0.4 – 1.2) | **0.182** | 0.8 (0.5 – 1.2) | 0.275 |
| **Level of education** |  |  |  |  |
| Nursery | 1 |  |  |  |
| Primary | 1.3 (0.5 – 3.3) | 0.623 | - | - |
| Secondary | 1.9 (0.7 – 5.3) | 0.195 | - | - |
| **Birth order** |  |  |  |  |
| First | 1 |  |  |  |
| Middle | 1.1 (0.4 – 1.4) | 0.741 | - | -- |
| Last | 0.7 (0.4 – 1.4) | 0.337 | - | - |
| **Person with whom the child resides (current custody)** |  |  |  |  |
| Both parents | 1 |  |  |  |
| Mother only | 0.7 (0.4 – 1.3) | 0.285 | - | - |
| Others | 1.5 (0.7 – 3.1) | 0.260 | - | - |
| **Missed any day during school term** |  |  |  |  |
| No | 1 |  | 1 |  |
| Yes | 2.5 (0.6 – 9.9) | **0.202** | 2.1 (0.5 – 8.6) | 0.321 |
| **Hospital visit in past 6 months** |  |  |  |  |
| No | 1 |  |  |  |
| Yes | 1.1 (0.3 – 4.1) | 0.869 | - | - |
| **Hospital admissions in past 6 months** |  |  |  |  |
| No | 1 |  | 1 |  |
| Yes | 1.6 (0.9 – 2.9) | **0.076** | 1.6 (0.9 -2.8) | 0.136 |

Note: cOR – Crude Odds ratio. aOR – Adjusted Odd Ration.

## DISCUSSION

In this cross-sectional study, we investigated the psychological distress of children with sickle cell disease in Mulago national referral Hospital, Kampala, Uganda. Our findings suggest that up to over 1 in 3 of the participants experienced psychological distress. Secondly, among the determined factors, age had a positive relationship with children aged 11 to 17 years almost 2-folds more likely to experience psychological distress compared to their counterparts aged 7 to 10 years while educational level, parental custody, birth order and number of hospital visits had no relationship with psychological distress in children and adolescents with SCA. These findings confirm the negative impact of SCD on the mental health of children in Mulago national referral Hospital, Kampala, Uganda.

In Africa, the psycho-social experience of children/adolescents living with sickle cell disease is poorly documented(14) and to the best of our knowledge, this is the first study describing it in Uganda. However, a cross-sectional study to establish the prevalence and factors associated with depression among adults with sickle cell disease at Mulago Hospital, Uganda reported uniquely higher proportion (68.2%) of adult patients with SCD suffering from depression (15). The prevalence in our study though being nearly half of this is not surprising as it further affirms that age has a positive relationship of psychological distress in SCD as documented in other studies. With interest in Sub-Saharan Africa, our results are congruent with those reported from a cross-sectional study among children/adolescent with SCD in Congo where 39.3% of the participants were found to have negative mental consequences (16) which is also supported with another study in Nigeria(17)

When compared to other international previous studies, our study findings of a high prevalence of psychological distress in children with SCD greatly co-relates with other study in USA which found up to 46% (18) and 48% in a similar study in Saudi Arabia (19).However, this finding of high prevalence of psychological distress in children is in contrast with the low prevalence noted by other researchers in the USA (20) and Saudi Arabia(21). Also, an earlier study conducted in the USA which evaluated depression in a group of children aged 8-15 with SCD reported no differences were found between children with SCD and healthy children (22) .Though data was collected at home when children were not acutely ill rather than in a hospital setting, and interviews at home may differ from those conducted in outpatient settings. Much as all studies recruited children and adolescents, the differences in their results may be methodological as their tools for assessment varied.

Psychological distress in children with SCD is associated with socio-demographic, clinical, therapeutic, and evolutionary factors which vary across studies. In this study, the main factor found to be associated with psychological distress in children with SCD is age with children aged 11 to 17 years almost 2-folds more likely to experience psychological distress compared to their counterparts aged 7 to 10 years. This is in congruent with a similar study in Nigeria which reported that depression in this population increased with increasing age (17). Generally, other studies argued though that adolescence has more depressive symptoms than younger children in the general population because of the peculiar stressors they experience (23). However, compared to the general population, adolescents with SCD still reports more depressive symptoms than younger children suggesting possibility of unique challenges among them.

In our study, nearly half of the participants felt they were inferior to other children, over a third felt people discriminated against them and they had bad luck while, nearly a quarter found it difficult to make friends which might probably explained the above as noted in another study (24)

This study had slightly more males, who have higher rates of psychological distress, however gender was non-predictive of psychological distress in this study. Similar to our study, previous researches reported absence of an association between gender and depression among children and adolescents with SCA (17,20, 21). An earlier study however believed that female sex is an important predictor of depression in SCA (25). The differences in the age group of recruited SCA patients may explained this controversy as adolescent girls are noted to have higher rates of depression than boys while pre-adolescence attenuate the gender differences.(26).

This study found that children with SCA had increasingly higher risk of developing psychological distress with increasing level of education. However, statistical significance was not achieved. This is congruent with a study which reported educational levels as non-significant predictors of depression (27). A similar study in Nigeria though noted a significant positive linear relationship between education level and depression scores of SCA children and adolescents (17).This finding however is contrasted by most previous studies which identified levels of formal education as predictors of depression symptoms as SCA patients with lower education showed increased risk of being depressed (19). Children with SCA might though be expected to have a low level of education because of school absenteeism and early school dropout due to clinical complications of the disease and deteriorating health (28).

### Study strengths and limitations of the study

The present study has provided new insight on the experiences of these children living with sickle cell diseases in Uganda and provides a basis for further research. However, these findings cannot be extrapolated to all children living with sickle cell disease as we sampled the children who came to the hospital in the study period. Additionally, our sampling procedure which was convenience sampling method could have missed the views of the children who were unable to come during the during the study period. Moreover, the questionnaire was self-administered which could have resulted in reporting and recall bias. Furthermore, the sample from the inpatient, outpatient units and different children were not equal and stratified which might have skewed the results.

## CONCLUSION

The scores for psychological distress in children with SCA in this study increased with increasing age. As children with SCA get older, on top of medical management, psychological compliment is needed in their routine care These findings suggest a need for targeted interventions and support systems for children in the 11 to 17 age group to address and mitigate psychological distress. This though, in a low resource setting like Uganda where there is limited availability of psychiatrist, calls for a need to train the primary care physicians, pediatric hematologist and even medical officers to provide such services. Additionally, the study highlights the importance of considering socio-demographic factors such as living arrangements and school attendance, in developing tailored strategies to promote mental well-being in this vulnerable population. Counselors and child psychologist may as well find this outcome useful as it could equip them with information that can be useful when communicated to the caregivers and parents of these children.

## Data Availability

All relevant data are within the manuscript and its Supporting Information files.

## FUNDING

Research reported in this publication was supported by the Fogarty International Center of the National Institutes of Health, U.S. Global AIDS Coordinator and Health Diplomacy(S/GAC), and Presidents Emergency Plan for AIDS Relief (PEPFAR) under Award Number 1R25TW011213. The content is solely the responsibility of the authors and does not necessarily represent the official views of the National Institutes of Health.

## ETHICAL DECLARATIONS

### Ethics approval and consent to participate

Our study received an exemption from the Institutional Review Board (IRB) review granted by the Mulago Research and Ethics Committee under protocol MHREC 2425 on 15^th^ February 2023 to run for a period of one (1) year. It solely utilized deidentified information. All activities were performed in line with the guidelines and regulations.

### Consent to participate

Written assent obtained from the child and consent from parent or guardian. Participating in the study was strictly voluntary in nature.

### Consent for publication

Not applicable

### Competing interests

The authors declare no competing interests.

### Role of funder/Sponsor

The funder had no role in the design and conduct of the study.

### Data sharing statement/Availability

The datasets used and/or analyzed during the current study available from the corresponding author on reasonable request.

### Author contributions

All authors made substantial contributions to conception and design, acquisition, analysis and interpretation of data. The authors equally took part in manuscript writing and gave final approval for the manuscript to be published in the current journal. The authors agree to be accountable for all aspects of the work.

## Notes

### Competing Interest Statement

The authors have declared no competing interest.

### Funding Statement

Yes

### Author Declarations

Our study received an approval from the Institutional Review Board (IRB) review granted by the Mulago Research and Ethics Committee under protocol MHREC 2425 on 15th February 2023 to run for a period of one (1) year.

## REFERENCES

1. Rees DC, Williams TN, Gladwin MT. Sickle-cell disease. Lancet. 2010 Dec 11;376(9757):2018–31.

2. Organization WH. Sickle cell disease prevention and control. Online at http://wwwafrowhoint/en/nigeria/nigeriapublications/1775-sickle. 2015;

3. Piel FB, Patil AP, Howes RE, Nyangiri OA, Gething PW, Dewi M, et al. Global epidemiology of sickle haemoglobin in neonates: a contemporary geostatistical model-based map and population estimates. The Lancet. 2013 Jan;381(9861):142–51.

4. Quinn CT, Rogers ZR, McCavit TL, Buchanan GR. Improved survival of children and adolescents with sickle cell disease. Blood. 2010 Apr 29;115(17):3447–52.

5. Ndeezi G, Kiyaga C, Hernandez AG, Munube D, Howard TA, Ssewanyana I, et al. Burden of sickle cell trait and disease in the Uganda Sickle Surveillance Study (US3): a cross-sectional study. Lancet Glob Health. 2016 Mar;4(3):e195–200.

6. Anie KA. Psychological complications in sickle cell disease. Br J Haematol. 2005 Jun;129(6):723–9.

7. Jerrell JM, Tripathi A, McIntyre RS. Prevalence and treatment of depression in children and adolescents with sickle cell disease: a retrospective cohort study. Prim Care Companion CNS Disord. 2011;13(2).

8. Hildenbrand AK, Barakat LP, Alderfer MA, Marsac ML. Coping and coping assistance among children with sickle cell disease and their parents. J Pediatr Hematol Oncol. 2015 Jan;37(1):25–34.

9. Levenson JL, McClish DK, Dahman BA, Bovbjerg VE, de A Citero V, Penberthy LT, et al. Depression and anxiety in adults with sickle cell disease: the PiSCES project. Psychosom Med. 2008 Feb;70(2):192–6.

10. Hasan SP, Hashmi S, Alhassen M, Lawson W, Castro O. Depression in sickle cell disease. J Natl Med Assoc. 2003 Jul;95(7):533–7.

11. Mubangizi V, Maling S, Obua C, Tsai AC. Prevalence and correlates of Alzheimer’s disease and related dementias in rural Uganda: cross-sectional, population-based study. BMC Geriatr [Internet]. 2020;20(1):48. Available from: 10.1186/s12877-020-1461-z

12. Everson SA, Maty SC, Lynch JW, Kaplan GA. Epidemiologic evidence for the relation between socioeconomic status and depression, obesity, and diabetes. J Psychosom Res. 2002 Oct;53(4):891– 5.

13. Batterham PJ, Werner-Seidler A, O’Dea B, Calear AL, Maston K, Mackinnon A, et al. Psychometric properties of the Distress Questionnaire-5 (DQ5) for measuring psychological distress in adolescents. J Psychiatr Res. 2024 Jan;169:58–63.

14. Lukoo RN, Ngiyulu RM, Mananga GL, Gini-Ehungu JL, Ekulu PM, Tshibassu PM, et al. Depression in children suffering from sickle cell anemia. J Pediatr Hematol Oncol. 2015 Jan;37(1):20–4.

15. Mubangizi V, Maling S, Obua C, Tsai AC. Prevalence and correlates of Alzheimer’s disease and related dementias in rural Uganda: cross-sectional, population-based study. BMC Geriatr. 2020 Feb 10;20(1):48.

16. Moyen E, Mpandzou GA, Boukoulou MJD, Diatewa JE, Batchi-Bouyou AL, Ossou-Nguié PM, et al. Psychological Experience of Children and Adolescents with Homozygous Sickle Cell Disease in Brazzaville. Open J Pediatr. 2021;11(01):35–49.

17. Ezenwosu O, Chukwu B, Ezenwosu I, Uwaezuoke N, Eke C, Udorah M, et al. Clinical depression in children and adolescents with sickle cell anaemia: influencing factors in a resource-limited setting. BMC Pediatr. 2021 Dec 1;21(1):533.

18. Jerrell JM, Tripathi A, McIntyre RS. Prevalence and treatment of depression in children and adolescents with sickle cell disease: a retrospective cohort study. Prim Care Companion CNS Disord. 2011;13(2).

19. Alhomoud MA, Gosadi IM, Wahbi HA. Depression among Sickle Cell Anemia Patients in the Eastern Province of Saudi Arabia. Saudi J Med Med Sci. 2018;6(1):8–12.

20. Graves JK, Hodge C, Jacob E. Depression, Anxiety, and Quality of Life In Children and Adolescents With Sickle Cell Disease. Pediatr Nurs. 2016;42(3):113–9, 144.

21. Sehlo MG, Kamfar HZ. Depression and quality of life in children with sickle cell disease: the effect of social support. BMC Psychiatry. 2015 Apr 11;15:78.

22. Noll RB, Reiter-Purtill J, Vannatta K, Gerhardt CA, Short A. Peer relationships and emotional well-being of children with sickle cell disease: a controlled replication. Child Neuropsychol. 2007 Mar;13(2):173–87.

23. Hankin BL, Mermelstein R, Roesch L. Sex differences in adolescent depression: stress exposure and reactivity models. Child Dev. 2007;78(1):279–95.

24. Hildenbrand AK, Barakat LP, Alderfer MA, Marsac ML. Coping and coping assistance among children with sickle cell disease and their parents. J Pediatr Hematol Oncol. 2015 Jan;37(1):25–34.

25. Hasan SP, Hashmi S, Alhassen M, Lawson W, Castro O. Depression in sickle cell disease. J Natl Med Assoc. 2003 Jul;95(7):533–7.

26. Friedberg RD, Sinderman SA. CDI Scores in Pediatric Psychiatric Inpatients: A Brief Retrospective Static Group Comparison. Depress Res Treat. 2011;2011:134179.

27. Alsubaie SS, Almathami MA, Abouelyazid A, Alqahtani MM. Prevalence of depression among adults with sickle cell disease in the southern region of Saudi Arabia. Pak J Med Sci. 2018;34(4):929–33.

28. Vilela RQB, Cavalcante JC, Cavalcante BF, Araújo DL, Lôbo M de M, Nunes FAT. Quality of life of individuals with sickle cell disease followed at referral centers in Alagoas, Brazil. Rev Bras Hematol Hemoter. 2012;34(6):442–6.

